# Adjuvant pembrolizumab after upfront multimodal therapy for stage IVB Anaplastic Thyroid Cancer

**DOI:** 10.1101/2025.03.22.25323852

**Authors:** Maria E. Cabanillas, Naifa L. Busaidy, Gary B. Gunn, Priyanka C. Iyer, Renata Ferrarotto, Maria Gule-Monroe, Anastasios Maniakas, Michelle D. Williams, Suyu Liu, Bryan Fellman, Michael Spiotto, Sarah Hamidi, Neal Akhave, Anna Lee, Jennifer R. Wang, Luana de Sousa, Vicente R. Marczyk, Mark Zafereo, Ramona Dadu

## Abstract

**Background:** ATC has historically been almost uniformly fatal. In patients with loco-regional disease (stage IVB), multimodal therapy (upfront surgery when feasible, radiation +/-concurrent chemotherapy) followed by observation is the current standard of care.

**Methods:** Stage IVB ATC patients treated with multimodal therapy, followed by adjuvant pembrolizumab were studied. Data were combined from a prospective, phase 2 trial (NCT05059470) that closed early due to poor accrual, and a retrospective cohort of consecutive patients who received adjuvant pembrolizumab and mirrored the trial eligibility criteria. Patients received adjuvant pembrolizumab starting within 6 weeks after completion of radiation. An age and treatment-matched control arm treated with multimodal therapy without adjuvant pembrolizumab was selected for comparison. The primary objectives included median progression-free survival (PFS) and recurrence rate. The secondary objective was median overall survival (OS). Descriptive statistics and Kaplan-Meier method for survival analysis were used.

**Results:** Between March 2020 and February 2024, 16 patients were treated with adjuvant pembrolizumab. The control arm included 16 patients. The median age in both groups was 59 years. The median PDL1 score and tumor mutation burden in the adjuvant pembrolizumab arm were 50% (range, 0-95%) and 3 mut/Mb (range, 0.5-29). There were more RAS mutated patient tumors in the adjuvant arm (40%) compared to the historical control group (18%). The majority, 14/16 (88%), had upfront surgery in both groups. The median follow-up time was 24.3 months in the adjuvant arm and 56.7 months in the control arm. The median PFS in the adjuvant and control arm was not reached, and 5.4 months (95%CI 2.04-16.20), respectively (p=0.006; HR 0.24 (95%CI: 0.08, 0.73)). The median OS was not reached in the adjuvant pembrolizumab group. In the control group the median OS was 31 months (95%CI 13.9, NA) (p = 0.009; HR 0.11 (95%CI: 0.01, 0.83)). The 12-and 24-month survivals were 80% (95%CI 0.51-0.93) and 52% (95%CI 0.25-0.74), respectively, in the control arm, whereas all patients in the adjuvant arm were still alive at 1- and 2-years.

**Conclusion:** Adjuvant pembrolizumab appears to be a safe and effective strategy to prevent recurrences and prolong survival in stage IVB ATC patients following multimodal therapy.

## Introduction

Until recently, a diagnosis of anaplastic thyroid cancer (ATC) was considered uniformly fatal. However, the past decade has ushered in a number of systemic therapies and treatment approaches which have shown efficacy in these patients (1-5). Patients with stage IVA (tumor confined to the thyroid) or IVB (loco-regional disease) have a far better prognosis than patients with distant metastatic disease (stage IVC) when treated with multimodal therapy. Traditional multimodal therapy consists of upfront surgery when feasible, radiation plus concurrent cytotoxic chemotherapy, followed by observation. This is the current standard of care included across all guidelines for ATC (6-8). However, relapse rates are high with multimodal treatment, likely due to microscopic metastatic disease, highlighting the need for effective adjuvant therapy. The Mayo Clinic group reported a 12-and 24-month overall survival (OS) of 68% and 48%, respectively, in this population. Relapses carry a very high risk of mortality, as 42% of patients died within 6 months (9).

In ATC, the PD-1 receptor-ligand interaction is a major pathway hijacked by tumors to suppress immune control and most ATC tumors express PD-L1 (10-12). Pembrolizumab, an anti-PD1 checkpoint inhibitor, blocks the interaction of PD1 and PDL1, resulting in an anti-tumor immune response. Thus, we hypothesized that adjuvant pembrolizumab would provide a local control benefit, in addition to targeting microscopic metastatic disease.

## Methods

Patients with stage IVB ATC who were undergoing or had completed upfront multimodal therapy were given consideration for a prospective, phase 2 trial with adjuvant pembrolizumab (NCT05059470), called IMPAACT (“ IMRT followed by pembrolizumab in the adjuvant setting in anaplastic cancer of the thyroid”). Unfortunately, the trial closed early due to poor accrual. Subsequently, a new study combining the data from 6 patients treated in the IMPAACT prospective trial with a retrospective cohort of consecutive stage IVB ATC patients who received adjuvant pembrolizumab outside of trial was created. An age- and treatment-matched historical group of patients with stage IVB ATC was selected as the control group. This study was performed in compliance with Good Clinical Practice guidelines and in accordance with the Declaration of Helsinki. The protocol and amendments were approved by the institutional IRB.

The retrospective cohort of patients treated with adjuvant pembrolizumab was chosen by using criteria that mirrored the trial eligibility criteria for IMPAACT. Eligibility criteria included: stage IVB ATC, completion of multimodal therapy (surgery when feasible and external beam radiation to the neck with or without concomitant cytotoxic chemotherapy), and adjuvant pembrolizumab within 6 weeks (+2 weeks allowed to account for latent toxicity resolution)) after the completion of radiation. As this was an intention-to-treat analysis, patients who were thought to have stage IVB disease due to small, FDG-non-avid equivocal lesions who were later identified as having metastases were included. Pembrolizumab dosing in the IMPAACT trial was 400mg IV every 6 weeks. Patients outside of trial could have received the 400mg IV every 6 weeks or 200mg IV every 3 weeks. For patients receiving infusions every three weeks, treatment cycles were defined as six-week periods, with each cycle consisting of two infusions. Patients in the clinical trial were planned to receive pembrolizumab for 1 year with an optional 2nd year per patient and physician discretion.

Patients in the historical control arm were selected from our IRB-approved ATC database. Those having received adjuvant immunotherapy during radiation or adjuvant treatment after radiation therapy were excluded. Patients were observed off all treatment after the completion of multimodal therapy, which is the current standard of care These patients were age-matched within 5 years of age, and treatment-matched by extent of surgery (R0/R1 vs R2) and dose of radiation (≥ 45 Gy vs < 45 Gy radiation). No patients prior to 2014 were selected.

The primary objectives of this trial were to estimate the recurrence rate and median progression-free survival (PFS) in stage IVB ATC patients treated with adjuvant pembrolizumab after the completion of initial standard therapy and compare them to the age and treatment-matched historical control group. PFS was determined by using the start date of pembrolizumab (for the adjuvant treated patients) or end of radiation (for the historical control patients), to the last radiographic assessment of progression or date of death. Secondary objective was to estimate and compare the median overall survival (OS) in the two cohorts, using the same starting dates as PFS.

RECIST v1.1 was used to evaluate relapses. For the efficacy endpoints, OS and PFS, the Kaplan-Meier method were used to estimate their survival functions. The marginal comparisons in PFS and OS were performed using the log-rank test. Cox regression model was used to estimate the hazard ratio (HR) of using adjuvant pembrolizumab, stratified by the matching factors of age and treatment.

## Results

### Treatment groups

A total of 16 patients between March 2020 and February 2024, were included in the adjuvant pembrolizumab group and an equal number of historical control patients between May 2014 and January 2024. Table 1 shows the baseline characteristics of both groups. Six (34%) of the adjuvant pembrolizumab patients were accrued in the IMPAACT trial, while 10 (63%) were identified from the retrospective study. Both arms were well balanced in terms of age, sex, and extent of surgery. One patient in each group did not undergo upfront surgery. All patients in the historical control arm received intensity modulated radiation (IMRT) ≥45 Gy and concomitant radiosensitizing chemotherapy. In the adjuvant pembrolizumab arm, all but one received IMRT ≥ 45 Gy with radiosensitizing chemotherapy. The patient who received lower dose radiation, underwent urgent palliative radiation (30 Gy in 10 fractions) using 3-dimensional conformal radiation to intact tumor, without radiosensitizing chemotherapy. The median dose of radiation between the two groups was similar.

**Table 1.** Baseline patient characteristics.

|  | Adjuvant<br>pembrolizumab arm<br>(N=16)<br>1/2020-1/2024 | Matched control arm<br>(N=16)<br>6/2014-3/2024 |
| --- | --- | --- |
| **Median age (range)** | 59 (47-76) | 59 (45-78) |
| **Sex, n (%)**<br>Female<br>Male | 10 (62)<br>6 (38) | 9 (56)<br>7 (44) |
| **Surgery**<br>R0/R1<br>R2 or biopsy only | 14 (87.5)<br>2 (12.5) | 14 (87.5)<br>2 (12.5) |
| **IMRT dose**<br>≥ 45 Gy<br>< 45 Gy | 15 (94)<br>1 (6) | 16 (100)<br>0 |
| **Median dose of RT (Gy)** | 60 | 66 |
| **Radiosensitizing**<br>**chemotherapy**<br>yes<br>no | 15 (94)<br>1 (6) | 16 (100)<br>0 |

### Mutation and PDL1 analysis

All patient tumors were sent for molecular analysis. Only 1 patient’s tumor (in the adjuvant pembrolizumab arm) could not be genotyped due to a very high immune infiltrate. Thus, 15/16 (94%) of the adjuvant arm tumors were evaluated by NGS for mutations and 13/16 (81%) evaluated for RNA fusions. In the matched historical control arm, all (100%) patient tumors were evaluated by NGS for mutations; 9/16 (56%) evaluated for RNA fusions. Figure 1 shows the oncoprint with patient characteristics and NGS data. In summary, there were more RAS mutations identified in the adjuvant pembrolizumab group (6/15, 40%) compared to the matched controls (3/16, 18%). Otherwise, there were similar numbers of driver oncofusions and mutations in BRAFV600E and PTEN. The median tumor mutation burden was 3 mut/Mb (range, 0.5-29) but there was insufficient aggregate information in the matched controls due to the historical nature of this group. The median PDL1 (Clone 22C3, Dako PharmDx) Tumor Proportion Score (TPS) was 50% (range, 0-95%) in the 15 patients whose tumors were tested in the adjuvant pembrolizumab arm. Insufficient aggregate data for PDL1 was available in the matched control arm.

**Figure 1.**
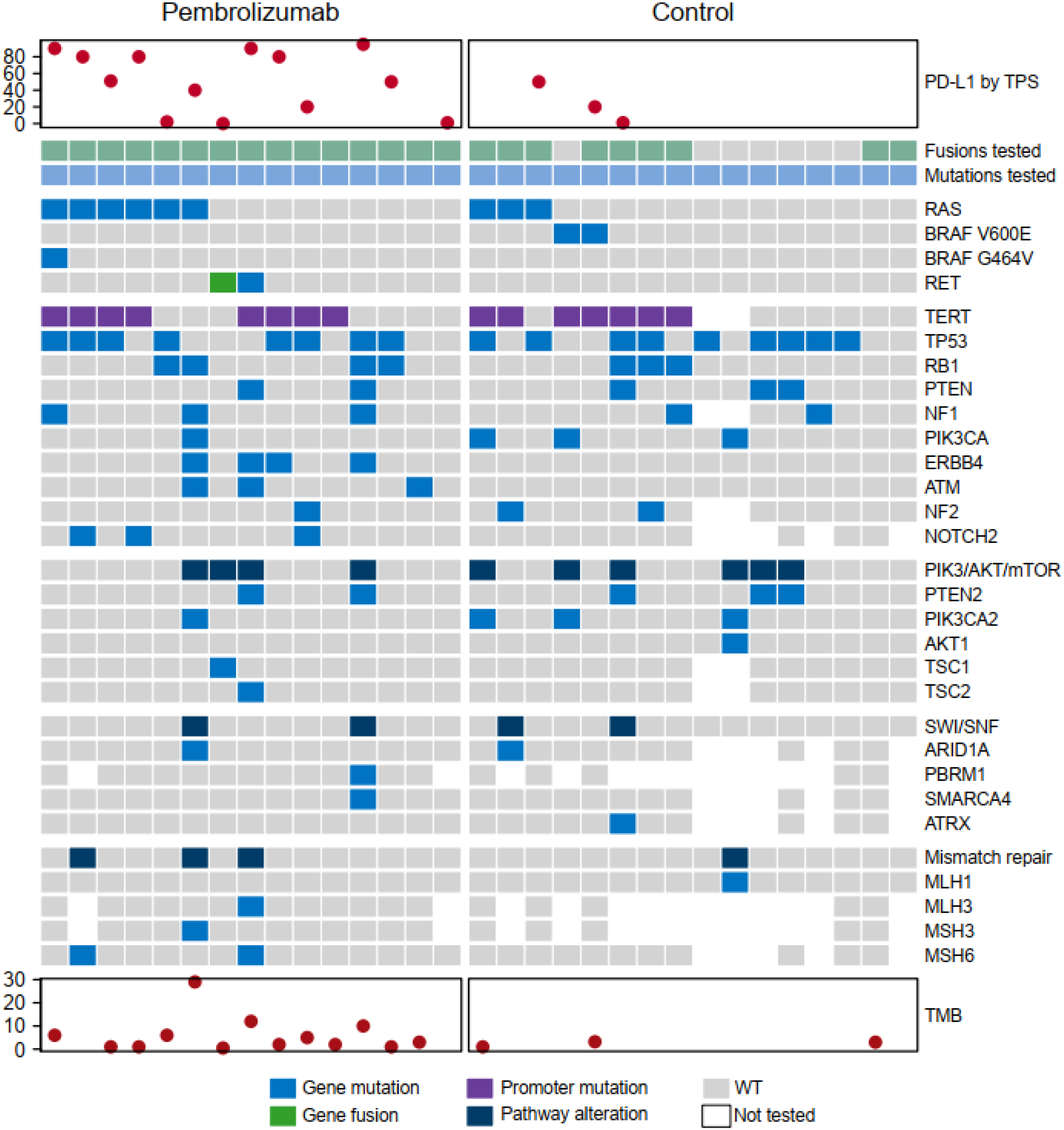
Oncoprint of targeted somatic gene mutations by Next-Generation Sequencing of tumors in patients with stage IVB Anaplastic Thyroid Cancer (ATC) in the adjuvant pembrolizumab and historical control arms. Several CLIA-certified panels were used to assay for mutations in oncogenes of ATC tumors, including liquid biopsy (shown in figure). One patient with BRAFV600E positive immunohistochemistry, who was in the adjuvant pembrolizumab arm, is not included in the oncoprint because mutation profile was not possible on his tumor.

### Efficacy results

Median follow-up time was 24.3 months for the adjuvant pembrolizumab group versus 56.7 months for the control arm. In the adjuvant pembrolizumab group, patients received a median of 16 cycles (normalized to 6-week cycles for the analysis; range 1-19 cycles) of pembrolizumab. At the data cut-off, only 2 patients were in active treatment with pembrolizumab. These two patients had completed 11 and 16 cycles of pembrolizumab.

There were 3/16 (19%) patients who were counted as having recurrence in the adjuvant pembrolizumab group and 14/16 (82%) in the matched control arm (p=<0.001). All 3 patients in the adjuvant pembrolizumab arm relapsed at distant sites, however in retrospect the metastatic lesions were all present at the time of starting pembrolizumab. These metastatic lesions were considered non-specific at the time of diagnosis and are discussed below. One patient treated with adjuvant pembrolizumab died after 2.74 years without having experienced progression. The median PFS was not reached in the adjuvant pembrolizumab arm but 5.4 months (95%CI 2.04, 16.20; p=0.006) in the matched control arm. The estimated HR was 0.24 (95%CI: 0.08, 0.73). The median OS was not reached in the adjuvant pembrolizumab groups. In the control group the median OS was 31 months (95%CI 13.9, NA; p = 0.009). The estimated hazard ratio was 0.11 (95%CI: 0.01, 0.83). At 1 and 2 years, all patients were alive in the adjuvant pembrolizumab arm, whereas 81% (95%CI 0.53, 0.95) and 54% (95%CI 0.27, 0.75) were alive in the matched control arm, respectively. Figure 2 shows the PFS (figure 2A) and OS (figure 2B) Kaplan-Meier curves, and the swimmer’s plot (figure 2C) of patients in the adjuvant pembrolizumab and historical control arms.

**Figure 2.**
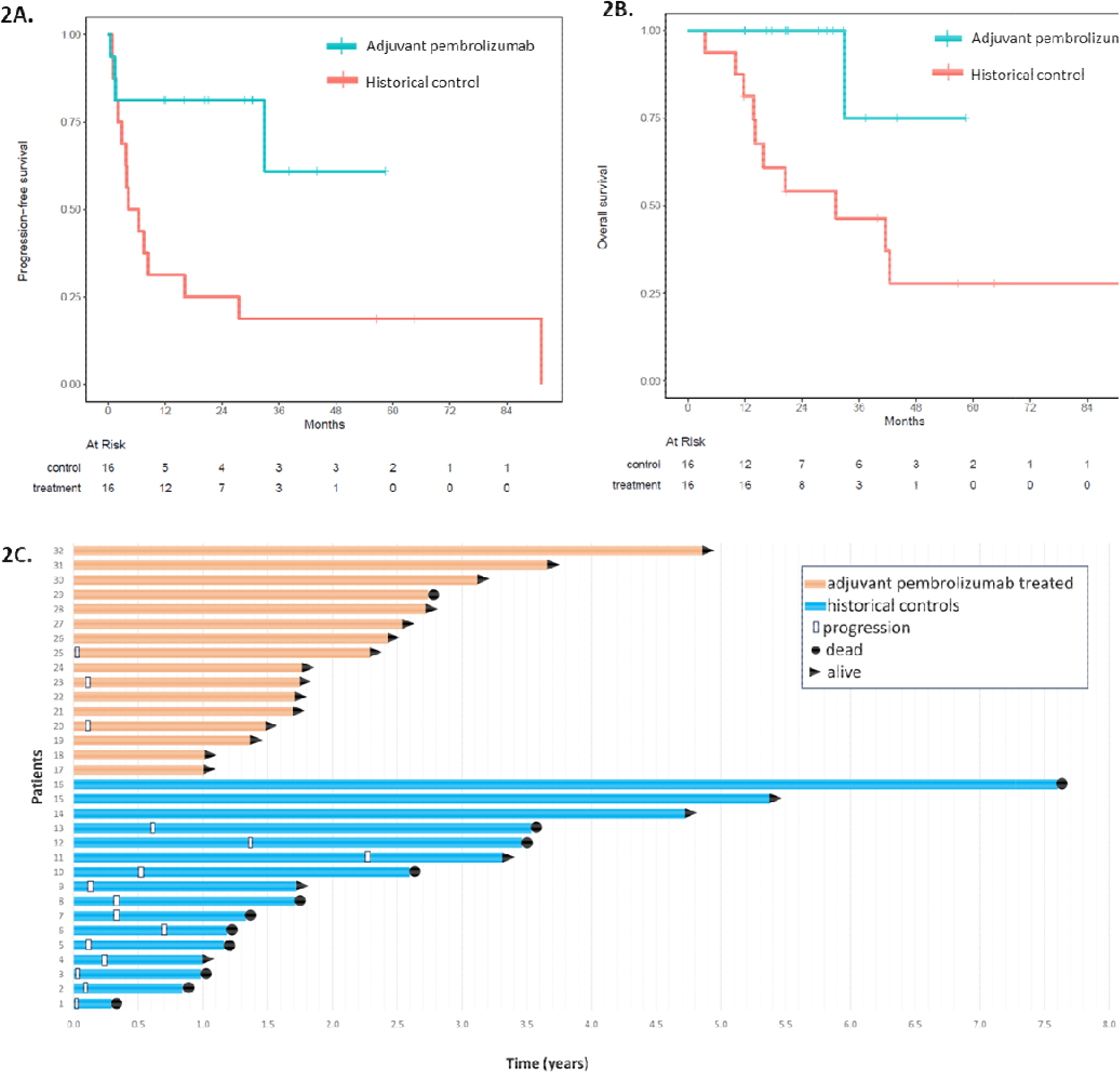
Median progression-free (2A) and overall survival (2B) in the adjuvant pembrolizumab cohort vs matched historical controls of stage IVB ATC patients. (2C) Swimmer’s plot showing the time in years from completion of radiation to progression and survival in stage IVB patients treated with adjuvant pembrolizumab or no adjuvant treatment. All patients, with the exception of patient 32, had upfront surgery prior to radiation. This patient had palliative radiation (30 Gy) to gross disease before starting adjuvant pembrolizumab.

In terms of serious adverse events (SAE) in the adjuvant pembrolizumab arm, grade 2 pneumonitis was reported in 1 patient. The patient was treated with steroids and pembrolizumab was permanently discontinued after 4 cycles. It was unclear if the SAE in the patient who died after developing gastric ischemia was related to pembrolizumab. The patient had chronic lymphocytic leukemia and was also taking acalabrutinib. He was admitted to an outside hospital with a diarrheal illness and found to have ischemic injury to his stomach leading to sepsis and death. No other serious adverse events were identified.

### Adjuvant patients who progressed

There were no true recurrences in the adjuvant arm. The 3 patients who were counted as having progression on adjuvant pembrolizumab were patients who had metastatic disease to the lungs prior to course 1 of adjuvant pembrolizumab. All three patients were alive at the data cut-off. Two of the three have required salvage therapy with targeted oral kinase inhibitors.

The first case is a man in his 50’s, status post total thyroidectomy, central and lateral neck dissection and excision of tumor thrombus from the left jugular vein (R0 resection) and radiation to the neck. His tumor’s PLD1 score was 20%, and the TMB was 5. He presented for the first cycle of adjuvant pembrolizumab in the clinical trial within 26 days after completing radiation. The baseline imaging showed growth of a small lung nodule which was present in a previous CT-scan but deemed indeterminate. This was biopsied and consistent with ATC. He proceeded with pembrolizumab, and at cycle 6 was found to have a complete response in the lungs. He completed 18 cycles of adjuvant pembrolizumab and then the treatment was stopped, as he had a complete response. He was alive without recurrence 27.8 months after starting pembrolizumab.

The second case was a woman in her late 70’s with RET fusion papillary thyroid cancer (PTC) diagnosed 22 years prior, which transformed into ATC. She underwent surgical resection with final surgical pathology showing ATC in background of PDTC in a lymph node. PDL1 was 0% and TMB was 0.5. She received radiation to the neck after surgery and started adjuvant pembrolizumab 47 days after completion of radiation. At baseline she had lung nodules, which then grew while she was on adjuvant pembrolizumab. These lung nodules were not biopsied nor confirmed to be ATC. She was removed from the clinical trial and started on a selective RET inhibitor with a partial response.

The third case was a man in his late 70’s with BRAFV600E mutated ATC who had an R2 surgery followed by radiation to the neck. PDL1, TMB and full molecular profile could not be performed due to high lymphocytic infiltrate in the tumor. The day he was scheduled to start adjuvant pembrolizumab outside of the trial, his CT chest showed new lung nodules. He proceeded with pembrolizumab, with the infusion starting 37 days after the completion of radiation. However, upon restaging, progression of the lung nodules was observed, and dabrafenib/trametinib were subsequently added to his immunotherapy. He achieved a complete response to triplet therapy.

## Discussion

This is the first clinical trial with adjuvant pembrolizumab for 1-2 years in stage IVB ATC. We found that the patients treated with adjuvant pembrolizumab within 4 weeks after completing multimodal therapy had a significantly longer median PFS and OS compared to the historical, age- and treatment-matched control patients who were observed after upfront multimodal therapy. The median PFS and OS have not been reached in the adjuvant pembrolizumab treated group with a median follow up time of 2 years. The only recurrences within the adjuvant pembrolizumab group were found to have been present prior to the first dose of adjuvant pembrolizumab. Even so, the recurrence rate was much lower in the adjuvant pembrolizumab patients, and these recurrences were ultimately deemed to be progressions of prior indeterminate lung lesions. Importantly, there were not locoregional recurrences/progression in the patients receiving adjuvant pembrolizumab.

There are several clinical trials in ATC showing that the outcome of patients with locoregionally confined diseases (M0) is far superior to those with distant metastatic disease (M1), however, all of these demonstrate that the vast majority of patients with locoregionally confined disease relapse or die within 1-2 years of their diagnosis, despite surgery and radiation (9, 13, 14). The NRG Oncology RTOG 0912 clinical trial with adjuvant pazopanib vs. placebo subanalysis showed in that patients with M0/MX status, median survival in the placebo arm was 8.6 months (95% CI 4.9, 16.2)(14). By the 2^nd^ year, only 4/21 (19%) patients in the placebo arm were still alive. However, these patients are not directly comparable to our historical control population due to differences in the surgical extent, which influences survival significantly. A retrospective study from MD Anderson showed that the median time to failure in stage IVB patients was 6.2 months and median OS was 12.3 months(13), however, only 41% had been treated with upfront surgery. Despite this, within 2 years almost all patients had relapsed and/or died.

A retrospective study from the Mayo Clinic of stage IVB patients from 2003-2015 with multimodal therapy showed that at 1 and 2 years, 68% and 48% were alive, respectively, compared to our current trial’s historical cohort at 80% and 52%, respectively. In the Mayo cohort 19/22 (86%) patients had relapsed within the first 2 years after completion of multimodal therapy. All but 2 of these patients relapsed at distant sites. At 5 years, 20/22 (91%) patients had relapsed. The difference between our historical cohort (31 months) and the Mayo cohort (22months) is possibly due to better salvage therapies in more recent years because we only selected patients who presented after 2014. Our patients would have had access to more modern treatments such as BRAF/MEK inhibitors, lenvatinib/pembrolizumab and clinical trials. This could explain the improvement in 1 year survival, however, at 2 years the MD Anderson and Mayo historical cohorts appear equivalent. This highlights the need for an adjuvant therapy to address micrometastatic disease. In our study, the patients treated with adjuvant pembrolizumab patients seem to have far better outcomes than both the MD Anderson and the Mayo Clinic historical cohorts, with no patients having true progression and only one death which was not due to ATC recurrence. Longer follow-up is needed to determine if disease recurrences will appear after 1-2 years of adjuvant pembrolizumab treatment. ATC patients likely need at least 5 years of follow up to understand whether the patients are cured.

All patients in the control arm and all but one in the adjuvant pembrolizumab arm had upfront surgery. More data is needed on outcomes of patients who receive palliative radiation (ie, < 45 Gy) without upfront surgery, followed by adjuvant pembrolizumab, as many patients do not undergo surgical resection either due to unresectable disease or unacceptable surgical morbidity. On the other hand, if adjuvant pembrolizumab after upfront multimodal therapy is found to be curative, it may be time to consider performing more radical surgeries in otherwise healthy patients with non-BRAF mutated stage IVB disease, similar to mucosal squamous cell carcinoma patients. Total laryngectomy has up to now largely been discouraged in these patients in the treatment guidelines for ATC. However, if a substantial number of stage IVB ATC patients are able to achieve long-term control with adjuvant pembrolizumab, then upfront extensive surgery (e.g. laryngectomy) becomes more acceptable in situations where checkpoint inhibitor therapy is possible.

Tumor driver mutation is important for determining prognosis in ATC patients. The presence of a somatic BRAFV600E mutation, which is actionable with BRAF/MEK inhibitor therapy, is associated with far better prognosis than RAS mutations (15). Since BRAFV600E mutated ATC patients usually undergo neoadjuvant BRAF/MEK inhibitor/anti-PD1 inhibitor, only 1 patient with BRAFV600E mutated ATC was included in the treatment arm. Thus, it is unclear if the strategy of adjuvant pembrolizumab alone is sufficient for the BRAFV600E mutated ATC patient. At this time, we continue to treat BRAFV600E mutated patients with neoadjuvant and adjuvant dabrafenib/trametinib/pembrolizumab (16). In our study, there were more patients with RAS mutated tumors in the adjuvant pembrolizumab arm, thus favoring the control arm since RAS mutations are associated with far worse outcomes than BRAF-mutant ATC (15). There was no difference in the number of patients with actionable mutations or fusions in the control versus adjuvant pembrolizumab groups. TMB and PDL1 scores were as expected in the adjuvant pembrolizumab arm but there was insufficient data in the control group, as many NGS panels did not include TMB, and testing for PDL1 score was not part of our standard practice prior to 2018.

The only serious adverse event that was clearly related to pembrolizumab group was pneumonitis, a known adverse effect of the checkpoint inhibitors. Immune-related adverse events associated with these drugs can be life-threatening and warrant careful follow-up prior to each infusion. Another patient died 4 months after the last pembrolizumab infusion due to gastric ischemia/sepsis but it is not clear if this was a related event.

Unfortunately, the prospective clinical trial, IMPAACT, closed early for poor accrual, therefore we pooled data from the IMPAACT trial and from consecutive ATC patients who were treated “ off-label” with adjuvant pembrolizumab. A number of factors could have contributed to the poor accrual to the trial. First, the trial was opened to enrollment in 2021, during the COVID-19 pandemic which severely slowed accrual. Second, pembrolizumab is commercially available in the United States and many physicians treated their patients outside of clinical trial for the convenience of the patient. Therefore, we combined data from the prospective clinical trial with real-world data in patients treated similarly. In order to determine whether the adjuvant treatment group benefitted from pembrolizumab, we created an age and treatment matched cohort. However, questions remain regarding efficacy of adjuvant pembrolizumab in patients with unresectable disease who undergo radiation to gross disease, as only one such patient was treated in our study. The patient has been alive and free of disease nearly 5 years. Other questions remain regarding adjuvant pembrolizumab treatment of patients with stage IVA disease.

The major limitation of this study includes the small number of patients; however, this is expected given the rarity of ATC. There is also a lead-time bias in the historical control group which could favor the adjuvant pembrolizumab group, as these patients were diagnosed more recently, and therefore have significantly less follow-up time. The control group could have worse outcomes due to being diagnosed in earlier years and there is likely a referral bias of patients who have relapsed. Even so, compared to data from other institutions, the overall survival of our historical controls is significantly longer at 31 months.

Based on the findings from this study we propose an update to the algorithm for patients with stage IVB ATC with BRAF wild-type disease (figure 3). Once a stage IVB patient is identified, a biopsy specimen should be rapidly analyzed to determine the BRAF status by immunohistochemistry or single gene BRAFV600E mutation testing. If BRAFV600E has been ruled out, we recommend surgery if an R0/R1 surgery can be accomplished, followed by external beam radiation to the neck with concomitant radiosensitizing chemotherapy. Palliative radiation can also be considered when full dose radiation is not possible. Adjuvant treatment should be started within 4-6 weeks of completing radiation since recurrences have been seen as early as 4 weeks in our historical cohort. Patients should be treated with 1-2 years of adjuvant pembrolizumab. Longer treatments are not advisable because of the risk of developing an immunotherapy-related serious adverse event, which can occur at any point during treatment with checkpoint inhibitor therapy. However, shortening treatment to 1 year could be studied in the future. Another area of future research is the use of adjuvant pembrolizumab in patients with stage IVA ATC, as well as those with BRAFV600E mutated tumors. Our trial did not include sufficient numbers of the latter. Because neoadjuvant therapy in BRAFV600E mutated ATC patients has been studied extensively, this is the preferred treatment for those patients. However, in countries where complicated surgeries are not possible or BRAF/MEK inhibitors are unavailable, adjuvant pembrolizumab could be considered for stage IVB BRAFV600E mutated ATC.

**Figure 3:**
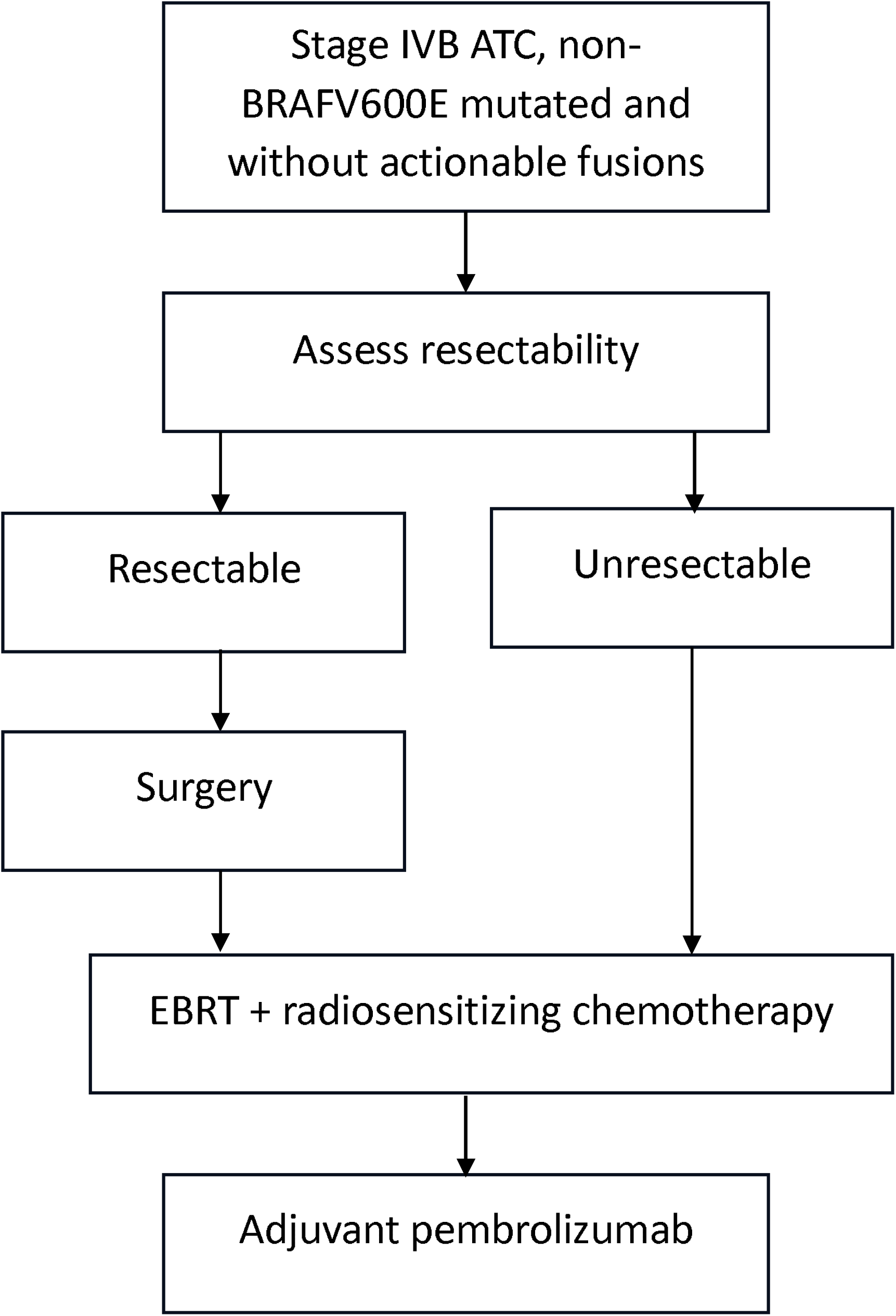
Proposed algorithm for patients with stage IVB anaplastic thyroid cancer without a BRAFV600E mutation. Once BRAFV600E has been ruled out, we recommend surgery when feasible, followed by external beam radiation to the neck with radiosensitizing chemotherapy. Palliative radiation may be considered when full dose radiation is not possible or acceptable. Patients should be restaged prior to adjuvant pembrolizumab. Ideally, adjuvant pembrolizumab should be started within 4-6 weeks for a period of 1-2 years. ATC=anaplastic thyroid cancer; EBRT=external beam radiation therapy

We conclude that adjuvant pembrolizumab appears to be a safe and effective strategy to prevent recurrences and prolong survival in stage IVB patients following upfront multimodal therapy and treatment guidelines should include this strategy.

## Data Availability

All data produced in the present study are available upon reasonable request to the authors

## Acknowledgements

We wish to thank Merck for providing drug support and the Petrick philanthropic funds for providing the funding for the trial. We also thank the brave patients and their families for their participation in the clinical trial and for entrusting their care to us.

## Author Contribution statement

Drafting of the manuscript: Maria E. Cabanillas. Conceptualization of the study: Naifa Busaidy, Gary B. Gunn, Renata Ferrarotto, Maria Gule-Monroe, Mark Zafereo, Ramona Dadu. Critical review and editing of the manuscript: All authors.

## References

1. Wang JR, Zafereo ME, Dadu R, Ferrarotto R, Busaidy NL, Lu C, et al. Complete Surgical Resection Following Neoadjuvant Dabrafenib Plus Trametinib in BRAF(V600E)-Mutated Anaplastic Thyroid Carcinoma. Thyroid. 2019;29(8):1036–43.

2. Zhao X, Wang JR, Dadu R, Busaidy NL, Xu L, Learned KO, et al. Surgery After BRAF-Directed Therapy Is Associated with Improved Survival in BRAF(V600E) Mutant Anaplastic Thyroid Cancer: A Single-Center Retrospective Cohort Study. Thyroid. 2023;33(4):484–91.

3. Subbiah V, Kreitman RJ, Wainberg ZA, Cho JY, Schellens JHM, Soria JC, et al. Efficacy of dabrafenib (D) and trametinib (T) in patients (pts) with BRAF V600E–mutated anaplastic thyroid cancer (ATC). J Clin Oncol 2017;35((suppl; abstr 6023)).

4. Sehgal K, Pappa T, Shin KY, Schiantarelli J, Liu M, Ricker C, et al. Dual Immune Checkpoint Inhibition in Patients With Aggressive Thyroid Carcinoma: A Phase 2 Nonrandomized Clinical Trial. JAMA Oncol. 2024;10(12):1663–71.

5. Cabanillas ME, Dadu R, Ferrarotto R, Gule-Monroe M, Liu S, Fellman B, et al. Anti-Programmed Death Ligand 1 Plus Targeted Therapy in Anaplastic Thyroid Carcinoma: A Nonrandomized Clinical Trial. JAMA Oncol. 2024.

6. Bible KC, Kebebew E, Brierley J, Brito JP, Cabanillas ME, Clark TJ, Jr., et al. 2021 American Thyroid Association Guidelines for Management of Patients with Anaplastic Thyroid Cancer. Thyroid. 2021;31(3):337–86.

7. Haddad RI, Bischoff L, Ball D, Bernet V, Blomain E, Busaidy NL, et al. Thyroid Carcinoma, Version 2.2022, NCCN Clinical Practice Guidelines in Oncology. J Natl Compr Canc Netw. 2022;20(8):925–51.

8. Filetti S, Durante C, Hartl DM, Leboulleux S, Locati LD, Newbold K, et al. ESMO Clinical Practice Guideline update on the use of systemic therapy in advanced thyroid cancer. Ann Oncol. 2022;33(7):674–84.

9. Prasongsook N, Kumar A, Chintakuntlawar AV, Foote RL, Kasperbauer J, Molina J, et al. Survival in Response to Multimodal Therapy in Anaplastic Thyroid Cancer. J Clin Endocrinol Metab. 2017;102(12):4506–14.

10. Chintakuntlawar AV, Rumilla KM, Smith CY, Jenkins SM, Foote RL, Kasperbauer JL, et al. Expression of PD-1 and PD-L1 in Anaplastic Thyroid Cancer Patients Treated With Multimodal Therapy: Results From a Retrospective Study. J Clin Endocrinol Metab. 2017;102(6):1943–50.

11. Bastman JJ, Serracino HS, Zhu Y, Koenig MR, Mateescu V, Sams SB, et al. Tumor-Infiltrating T Cells and the PD-1 Checkpoint Pathway in Advanced Differentiated and Anaplastic Thyroid Cancer. J Clin Endocrinol Metab. 2016;101(7):2863–73.

12. Zhu X, Hu C, Zhang Z, Zhu Y, Liu W, Zheng B, et al. PD-L1 and B7-H3 are Effective Prognostic Factors and Potential Therapeutic Targets for High-Risk Thyroid Cancer. Endocr Pathol. 2024;35(3):230–44.

13. Rao SN, Zafereo M, Dadu R, Busaidy NL, Hess K, Cote GJ, et al. Patterns of Treatment Failure in Anaplastic Thyroid Carcinoma. Thyroid. 2017;27(5):672–81.

14. Sherman EJ, Harris J, Bible KC, Xia P, Ghossein RA, Chung CH, et al. Radiotherapy and paclitaxel plus pazopanib or placebo in anaplastic thyroid cancer (NRG/RTOG 0912): a randomised, double-blind, placebo-controlled, multicentre, phase 2 trial. Lancet Oncol. 2023;24(2):175–86.

15. Wang JR, Montierth M, Xu L, Goswami M, Zhao X, Cote G, et al. Impact of Somatic Mutations on Survival Outcomes in Patients With Anaplastic Thyroid Carcinoma. JCO Precis Oncol. 2022;6:e2100504.

16. Hamidi S, Dadu R, Zafereo ME, Ferrarotto R, Wang JR, Maniakas A, et al. Initial Management of BRAF V600E-Variant Anaplastic Thyroid Cancer: The FAST Multidisciplinary Group Consensus Statement. JAMA Oncol. 2024;10(9):1264–71.

